# Reducing Covid-19 risk in schools: a qualitative examination of staff and family views and concerns

**DOI:** 10.1101/2020.10.25.20216937

**Authors:** Ava Lorenc, Joanna M Kesten, Judi Kidger, Rebecca Langford, Jeremy Horwood

## Abstract

**Background:** The Covid-19 pandemic has necessitated schools implementing Covid-19 risk-reduction measures.

**Methods:** We investigated young people, parent and school staff attitudes towards secondary school Covid-19 mitigation measures. Recruitment used school communication, community organisations and snowball sampling. Audio recorded online/phone individual/group interviews lasted 45 minutes. Interviews focused on social distancing, hand-hygiene and testing. Team framework analysis used interview notes and transcripts.

**Results:** Participants were 13 school staff, 20 parents and 17 young people. Concerns about Covid-19 risk at school, especially to vulnerable individuals, were outweighed by perceived risks of not returning to school. Some teachers anticipated guilt around being a potential ‘spreader’. Participants saw school mitigation measures as an acceptable and pragmatic solution to the impossibility of social distancing, although anticipated challenges in changing habitual behaviour. Participants supported school Covid-19 testing but identified the need to consider data security and stigma. Staff were concerned about unintended consequences of risk-reduction strategies and widening inequalities.

**Conclusion:** Families and staff supported Covid-19 mitigation measures in schools. Clear messaging and engendering collective responsibility are important for compliance and success. However, schools and policy makers should consider unintended consequences of measures, supporting vulnerable individuals and those with additional needs, and avoiding widening inequalities.

## Introduction

To help reduce the rapid spread of Covid-19, UK ‘lockdown’ was announced on 23rd March 2020 (1) and school campuses were closed to all but a small group of vulnerable or priority students (2). In July, the government announced all school campuses would fully reopen in September (3).

Evidence for school closures reducing infection spread is equivocal (4). Those in favour of school campuses reopening to all pointed to the impact on learning (5), particularly for those with lower socioeconomic status (6), widening inequalities and the impact on students’ physical and mental health (7-14). Remote learning also impacted staff wellbeing and mental health (15). However, schools reopening carried risks - modelling suggested a second UK wave of Covid-19 infection would occur if schools reopened full time (combined with accompanied easing of restrictions in society and without scaled up test and trace systems) (16).

Government guidance released over the summer suggested a range of protective measures in schools to reduce the risk of Covid-19 outbreaks (18), including hand-hygiene, cleaning, reporting symptoms, and social distancing (3). Further, a UK modelling study recommended existing testing should be scaled up to avoid a second wave of Covid-19 when school campuses fully reopened (16, 17).

Understanding the views and concerns of school staff, parents and students about school Covid-19 measures being introduced (17) is key to ensuring compliance and avoiding unintended harms. This secondary school qualitative study aimed to rapidly explore young people, parent/carer and school staff attitudes towards school Covid-19 mitigation measures, views on managing Covid-19 infections in schools and opinions about student groups who may be particularly affected by these measures.

## Methods

### Study Setting

Between July and September 2020, we recruited 11-16 year olds, parents/carers and school staff through secondary schools and local community organisations in the South West of England.

### Sampling and recruitment

Because lower socioeconomic and Black, Asian and Minority Ethnic (BAME) populations are disproportionately affected by Covid-19, we contacted 21 schools with higher levels of these populations to ensure we captured their specific concerns. Participating schools sent study information to students/parents/carers/school staff. We focused student/parent/carer recruitment on Year 8 (age 12-13; most had been out of school since March) and Year 10 (age 14-15; faced key exams next year). Recruitment strategies also included advertising the study via community organisations (through their newsletters, social media, and direct contact with members) and snowball sampling via participants. Individuals interested in participating contacted the researcher directly to arrange an interview. All eligible volunteers were interviewed (none refused or dropped out).

### Interviews

Participants chose to be interviewed on their own, with a friend and/or parent/carer (young people) or with colleagues (staff). AL led interviews by phone or video using a secure internet platform, with either JMK or JK present for group interviews/child interviews without a parent present. Interviews were 45minutes and digitally audio recorded. Prior to the interview, participants provided audio-recorded verbal informed consent (or assent and parent/carer consent for those under 16 years).

Young people and school staff provided input into the study design and topic guides via two digital webinars.

Topic guides covering attitudes towards social distancing, school hand-hygiene and infection control strategies, and acceptability of a test and trace system were used flexibly, allowing exploration of issues raised by the participant. As thanks for their time, participants were given a shopping voucher after their interview.

Data collection was informed both by the concept of information power (18) and pragmatic considerations of the project timeline.

### Analysis

Producing timely reports (19, 20) for local and national stakeholders necessitated rapid analysis. AL used the framework method (21) to analyse anonymised notes taken during interviews, and re-listening to aspects of the interviews. Notes were coded deductively using topic guide headings to develop a coding framework in Microsoft EXCEL and identify themes. Framework analysis was appropriate for the specific a priori questions and limited time frame. Participants did not check the coding.

Audio recordings were transcribed and anonymised. The research team analysed the transcripts collaboratively, reading the transcripts and adding new subthemes and direct quotes to the existing framework.

The study was approved by the Faculty of Health Sciences Committee for Research Ethics, University of Bristol (ref 108084).

## Results

### Participants

Participants were 13 school staff (Heads/assistant Heads, teachers, Special Educational Needs Coordinators (SENCOs)) from seven schools - and 20 families from ten schools (12 with a BAME child/parent) - 17 young people (mostly years 7/8/10; 9 girls; 8 boys) and 20 parents (19 mothers, 1 father) (Table 1).

**Table 1.**
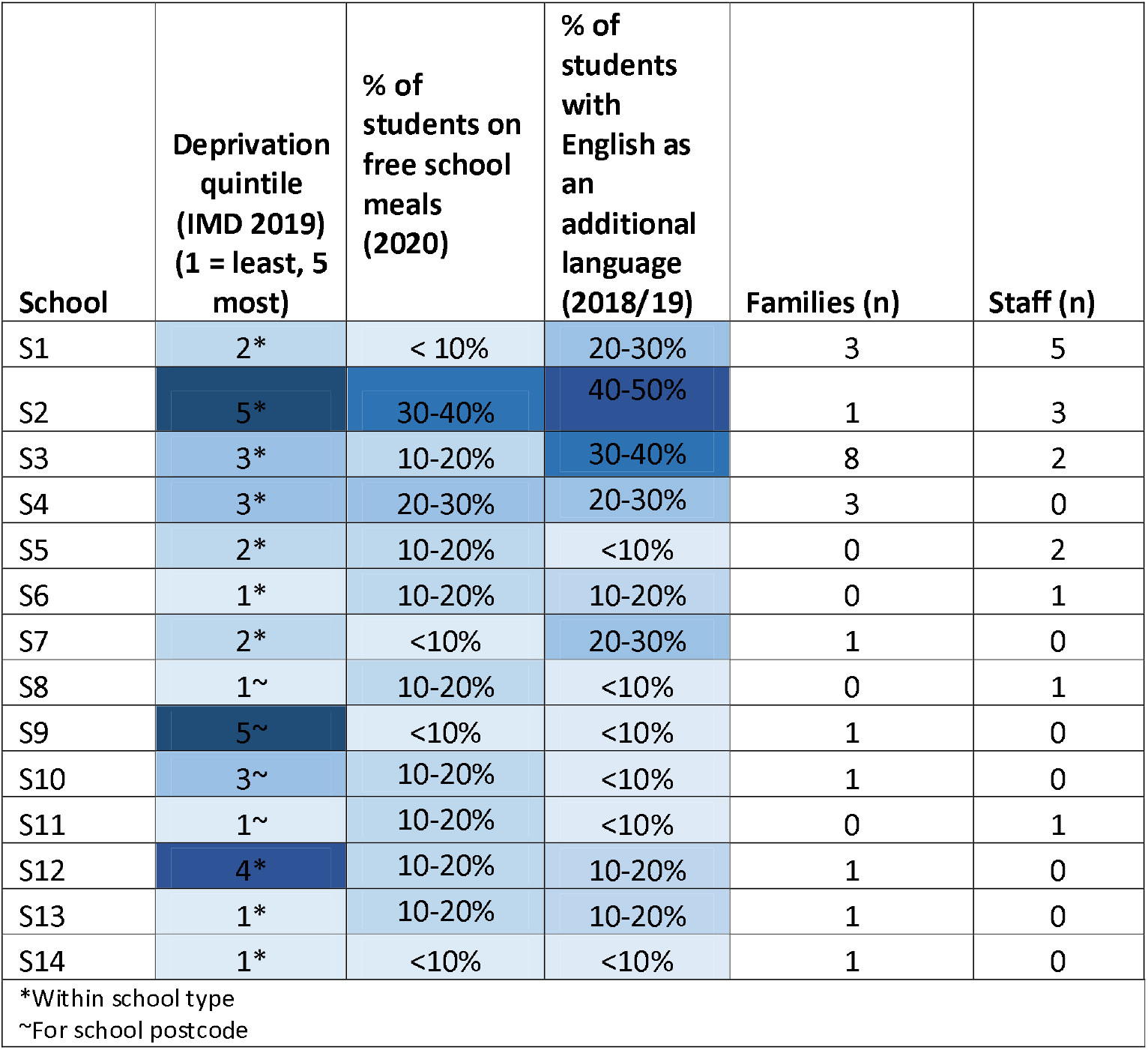
Participant characteristics.

### Covid-19 risk concerns

Most staff and around half of families (young people and parents) were not concerned about personal/family Covid-19 risk from returning to school. However, many staff anticipated increased Covid-19 cases – describing schools as “petri dishes”. Some staff worried about being a potential ‘spreader’ (Table 2). Staff also had concerns about higher risk staff and students, including those from BAME backgrounds, those who were pregnant or had health conditions.

**Table 2:**
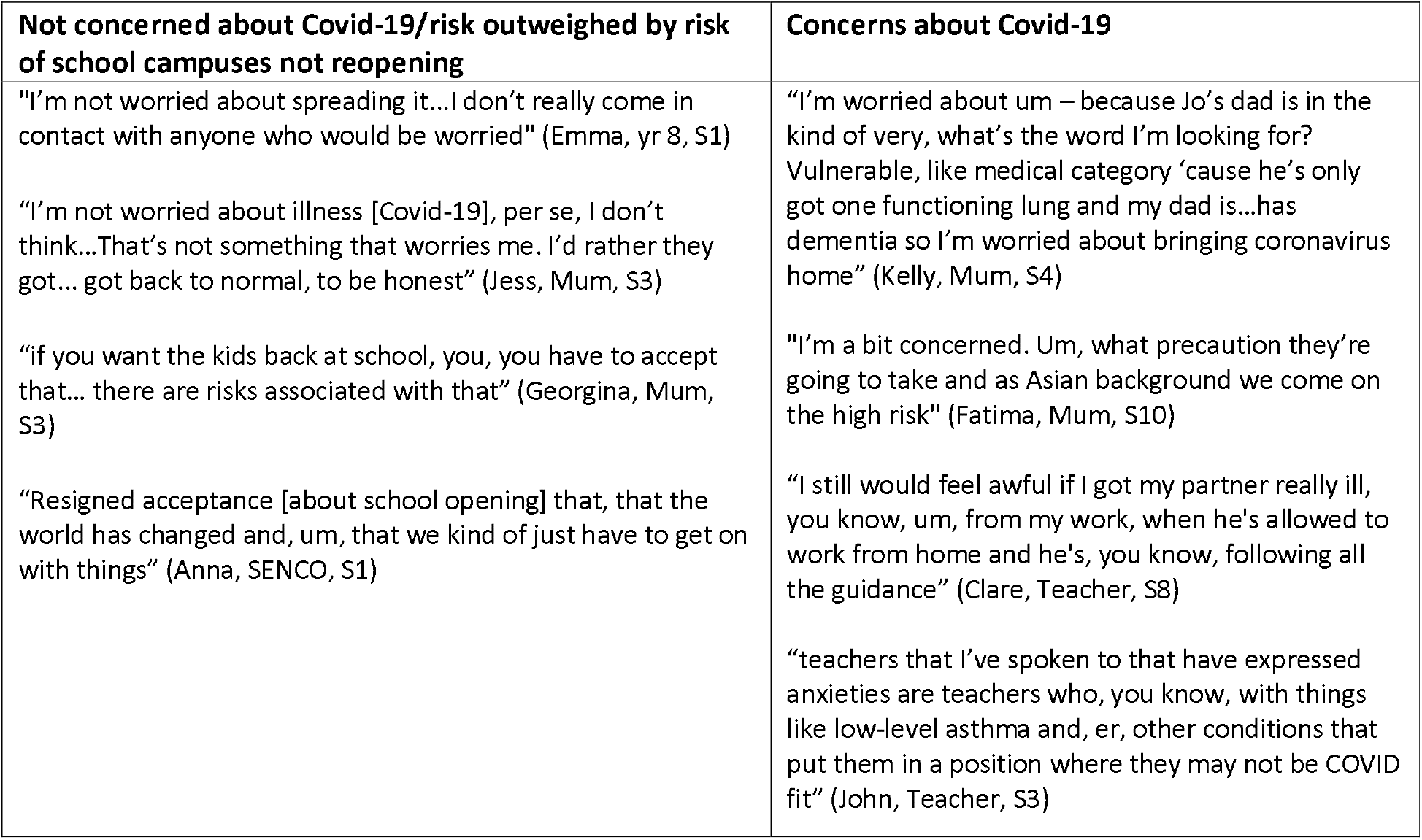
Quotes on concern about risk of Covid-19.

| **Not concerned about Covid-19/risk outweighed by risk of school campuses not reopening** | **Concerns about Covid-19** |
| --- | --- |
| "I'm not worried about spreading it...I don't really come in contact with anyone who would be worried" (Emma, yr 8, S1)<br>"I'm not worried about illness [Covid-19], per se, I don't think...That's not something that worries me. I'd rather they got... got back to normal, to be honest" (Jess, Mum, S3)<br>"if you want the kids back at school, you, you have to accept that... there are risks associated with that" (Georgina, Mum, S3)<br>"Resigned acceptance [about school opening] that, that the world has changed and, um, that we kind of just have to get on with things" (Anna, SENCO, S1) | "I'm worried about um – because Jo's dad is in the kind of very, what's the word I'm looking for? Vulnerable, like medical category 'cause he's only got one functioning lung and my dad is...has dementia so I'm worried about bringing coronavirus home" (Kelly, Mum, S4)<br>"I'm a bit concerned. Um, what precaution they're going to take and as Asian background we come on the high risk" (Fatima, Mum, S10)<br>"I still would feel awful if I got my partner really ill, you know, um, from my work, when he's allowed to work from home and he's, you know, following all the guidance" (Clare, Teacher, S8)<br>"teachers that I've spoken to that have expressed anxieties are teachers who, you know, with things like low-level asthma and, er, other conditions that put them in a position where they may not be COVID fit" (John, Teacher, S3) |

Many families were more concerned about the negative consequences of young people not being at school (e.g. impact on education) than Covid-19 risk, accepting that home schooling cannot continue indefinitely. However, families did note the risk to vulnerable family members (e.g. due to health or age). Concern was more common in families with BAME members, although only a minority explicitly cited ethnicity-related risk, with some noting the lack of scientific understanding around this risk.

### Covid-19 school risk reduction measures

Outside school, most young people had accepted social distancing, wearing masks and handwashing, understanding the rules and their necessity, with minor negative comments such as social distancing being ‘not nice’. Most aimed to social distance and a few had socialised very little and enjoyed staying at home.

Box 1 outlines the commonest school Covid-19 risk reduction measures identified by interviewees. Participants agreed on the need for such measures, in line with national Covid-19 guidance, although staff were frustrated with the lack of detail in this guidance, meaning each school had to develop their own plans. Staff were generally happy with schools’ plans but had reservations about feasibility and how they would “play out in practice”.

#### Barriers/concerns

All participants agreed student social distancing was impossible and “pointless”, given the numbers of people and lack of space (table 3). Year ‘bubbles’ (see Box 1) were seen as a “pragmatic solution”, although there was concern about crossover via siblings or groups leaving school together, and teachers (though less commonly reported). In terms of compliance with new risk reduction measures, of most concern was forgetting, and a desire to “be normal”.

**Table 3:** Quotes about school covid-19 risk reduction measures.

| Concerns/barriers | Facilitators |
| --- | --- |
| Social distancing impossible | Common good |
| "I can't see it [social distancing] happening unless there are less children in the school at one time" (Sarah, Mum, S4)<br>"you can't properly socially distance in a school so it's a lot of 'that'll do, that'll do' but it's still best it can be" (Matt, Teacher, S3)<br>"maybe they [school] have some magical plan, or maybe if they can extend something or something like that, but I... I just feel like there's not enough space for what they're trying to do" (Amber, yr 7, S3) | "I think empowering people to be autonomous and realise that their efforts are, are for the collective good. That they've all got a part to play in it. I think that's much better than, you know, er, publicly shaming them, or or, or criticising them....if you want to achieve compliance and cooperation, it's creating an atmosphere that it's in everyone's best interest and you contribute to the common good" (John, Teacher, S3) |
| Concerns about year bubbles | Messaging and reminders |
| "at the end of the day, we're all gonna walk out of school at the same time and we're all just gonna walk together and end up in the same place " (Amber, yr 7, S3)<br>"siblings they are mixing bubbles, the bubbles become 500, 700 [people], um, so it's a gesture" (Claire, Teacher, S8) | "if there's a consistent message, 'this is just what you do now, if you can', then, um, then they'll just pick it up really and get on with it" (Georgina, Mum, S3)<br>"I think having more visuals is really quite key so there's, they are reminded of what's, what's going on. Um, I don't feel like we have enough at the moment, to be honest" (Julia, Teacher, S11) |
| Concerns about hand-hygiene | Staff culture shift |
| "I suspect we're going to run out of hand sanitisers in the, um, the ones that are kind of communally put up around the school ...I'd say that they, they're going to run out by break time" (Julia, Teacher, S11)<br>"They [school bathrooms] never have soap or anything, like, loads of the taps don't work!" (Daisy, yr 7, S3) | "our biggest weapon will be 'don't be in work if you're ill'" (Jackie, Head, S1) |
| Lack of compliance with new rules |  |
| "[on handwashing] maybe forgetting or teachers not reminding the students enough or them being too lazy or maybe their friends might be going somewhere but their friends don't wanna wait for them so they just can't be bothered to wash their hands" (Ali, yr 10, S13)<br>"I feel like, you know, people are just are not gonna do it [social distance]. They're just gonna do what they want and like run around the whole place " (Amber, yr 7, S3)<br>"a line that's been put on the floor that shows the two metres to the teacher. Um, what's interesting there, of course some students are going to see that as a bit of an invite or a challenge to cross that line, literally" (Tracey, Mum, S3)<br>"[my child] might just go to the sink, wash the few fingers at the front and then leave. We don't actually know if they're going to be doing the full happy birthday... it baffles me how it's [enforcing handwashing] actually going to work" (Samira, Mum, S4) |  |
| Specific groups |  |
| "disadvantaged students, and students that would normally get |  |
| more support in a lesson - because it's more like lecture style now, so we're not really allowed to walk between the desks or anything - those students are going to miss out, the ones who wouldn't normally put their hand up " (Finn, Teacher, S5)<br>"the young people ... with the highest level of needs, who've got education health and care plans ...might not fully understand the need for personal space, or...we've just added this whole extra kind of layer of complexity into their social world that I'm sure is going to be hugely challenging" (Anna SENCO, S1) |  |

Staff and family concerns about hand-hygiene/infection control were mostly practical, including: availability of resources (sanitiser/soap, sinks, cleaners) - one school estimated a £40k cost of hand sanitiser; cleanliness of bathrooms; lack of time for handwashing; and effective use of measures e.g. hand sanitiser vs washing, or proper use of masks. Some were concerned about ventilation e.g. windows not opening.

A minority of staff were worried about behavioural issues arising from students having to stay in the same classroom e.g. unsupervised lesson changeovers, student boredom and lack of movement. Other concerns included Covid-19 risk of using public transport and a reduced range of lessons/activities. An important concern for staff was the impact of the risk reduction measures on learning and pastoral care, especially social distancing measures, e.g. fewer interactive lessons and less opportunity to provide extra support to individual students.

Staff were concerned social distancing measures would particularly affect students with special educational needs (SEN) or mental health issues. Particular issues for young people with SEN included (Table 3): struggling to understand the rationale for and to comply with the changes; finding less interactive lessons challenging; SEN support staff being unable to work closely with students; physical needs e.g. personal care or feeding; and removal of ‘safe spaces’ (due to bubbles and infection risk from soft furnishing).

#### Facilitators/suggestions

The main suggestion from staff for facilitating the new rules was educating young people about their importance, and encouraging a “we culture” of collective responsibility through a supportive, considerate approach (Table 3), although, a minority of families thought handwashing rules should be compulsory and enforced.

Staff and parents also suggested clear consistent messaging and daily reminders, both verbal and visual. Clear and regular communication from schools about the measures would also reassure parents (Table 3).

Other suggestions to support the mitigation measures included:

- Funding for cleaning products, PPE and hand sanitiser; additional classroom equipment to ensure no sharing
- Government guidance on PPE and SEN
- Staff training on how to work within the new measures
- Risk assessments for vulnerable students/staff
- Shorter lessons to allow time for handwashing
- Young people bringing their own hand sanitiser
- Encouraging staff to not come to school if displaying symptoms (a culture shift from presenteeism common in school staff) (Table 3).

### School management of Covid-19 cases

#### School response to Covid-19 cases

Several families anticipated possible stigma around Covid-19 diagnosis. Staff did not generally anticipate stigma, due to acceptance of Covid-19, and existing school population diversity generally promoting tolerance - although two mentioned possible “mass hysteria” and another that teenagers “love to joke and point a finger”.

> “if someone coughed in my class, I would see one or two people shying away from them a bit and that most of the class is laughing a little bit and saying, ‘oh you have Corona virus’ as a joke” (David, yr 8, S3)
>
> “I think everyone would just be very pragmatic about it and I can’t see there being panic stations” (John, Teacher, S3)

#### Reporting symptoms

Most families and some staff anticipated under-reporting of student Covid-19 symptoms, due to embarrassment, wanting to attend school, or parents needing to work. For staff, underreporting reasons included a culture of presenteeism, and guilt at having been in a risky situation. Families wanted clarity about symptoms to report, method of reporting and implications.

> “I think it will be very tricky… for parents to differentiate… fever and cough, they are quite common symptoms kids get in the winter season” (Sangita, Mum, S13)

#### Test and trace

All staff and most parents thought testing in schools was important and would be enthusiastic about monthly testing. Testing at schools would reassure students, parents and staff about school safety, and encourage attendance - a “massive selling point for schools”.

> “If there was a risk we were going to get the virus and it would make everyone safer then I would do it [regular testing]” (Jasmine, yr 10, S4)

A minority of participants had concerns about a school test and trace system, including:

- Parental concerns about use and anonymity of information about their child, particularly among BAME families “I understand the need [to collect data] but on the same token, it’s just that personal data being collected about my child makes me feel very uncomfortable” (Sarah, mum, S4)
- More school closures due to cases detected by testing, with implications for parents (time off work), young people (loss of learning) and schools (attendance figures and academic achievement).
- Feasibility of testing the whole school – time, space and administration needed, and challenges related to year group ‘bubbles’.

Similar to risk reduction measures, staff suggested emphasising to families the importance of testing, including a potentially reduced risk of whole school closure. They also emphasised the need for quick and minimally disruptive testing. Families wanted care, discretion and anonymity in notification of positive results.

> “market it towards the fact that they’ve missed so much education, this is something that can potentially help keep them in school for as long as possible. Because that’s ultimately what I think most parents are concerned about” (Dan, head of yr 8, S2)

## Discussion

### Main findings

Some families and school staff had concerns about an increased Covid-19 risk with the reopening of school campuses, particularly to vulnerable individuals, but on balance the majority felt the benefits outweighed the risks. Some staff anticipated guilt at their potential to spread Covid-19. Young people, their parents and school staff generally felt planned school risk reduction measures (see Box 1) were acceptable and pragmatic solutions to the perceived impossibility of social distancing in crowded schools.

Negative unintended consequences of the new measures were anticipated, on student behaviour, learning, pastoral care, and particularly for those with SEN or mental health issues who may find the measures especially challenging.

Stigma related to Covid-19 positivity was thought unlikely to be widespread in schools but the risk should be considered in plans for how case reporting is managed. The imperative for testing in schools was recognised by staff and most parents, albeit with some concerns over data security and feasibility.

### What is already known on this topic

UK teachers are concerned about the impact of the pandemic on learning, pastoral care and student wellbeing, especially in deprived communities, and the difficulties schools face in dealing with this (9, 22)(9, 22)(9, 22)(9, 22). Young people may be more affected by social distancing measures than other age groups (23). Staff and parents are particularly concerned about SEN students (24, 25). Evidence as to whether these concerns are justified is not yet available, but reports exist of worsening mental health among children(26), especially those from BAME backgrounds(27), and worsening student behaviour, partly due to risk reduction measures, including among SEN students (28). A compassionate ethos can address anticipated emotional and mental health needs of students(9). Frontline workers, which includes teachers now school campuses are fully re-opened, currently have high levels of anxiety and depression and distress, partly related to feelings of guilt and conflicting duties (29, 30).

Some data suggests parents/carers are concerned about the impact on their child’s mental health of mitigation measures (31) or that their child will not understand the need for social distancing (25). Data from lockdown and previous pandemics raises concerns about adherence and practical feasibility (32, 33) e.g. handwashing vs sanitiser(34) and social distancing challenges in secondary schools due to individual timetables and specialist equipment needs(35). To increase adherence, school messaging should educate and enable, emphasising benefits for family or wider community, and a sense of collective identity and responsibility(36-38), although ‘enlisting’ young people is not without challenges (39). Information needs to be clear, consistent, transparent and detailed (24, 40, 41), precisely defining required behaviour(36), and making things easy (42). Schools will also need to use policies, rules and reward systems, and discipline where needed (28, 36).

School leaders have called for funding and support to implement risk reduction measures (including supplies, self-care and extra staff to cover absences) (22, 36, 43). Schools reportedly spent up to £8k each on Covid-19 risk reduction measures in early reopening of campuses(44). Scientists also recommended funding to allow for smaller socially distanced classes (45)

The importance of test and trace programmes to prevent Covid-19 outbreaks in schools is clear (46-48), but current testing capacity in the UK is insufficient(41). There is little data on family and school views about testing, but public engagement work around Covid-19 vaccine trials found general apprehension, scepticism and low levels of trust in such research among in BAME communities (49). There is evidence of stigma associated with Covid-19, but not specifically from schools (50).

### What this study adds

Staff and families supported the UK government pledge to keeping school campuses open, and, contrasting with previous data (25), we found young people, parents and staff did understand the need for risk reduction measures, and generally reported compliance. However, effectiveness may be reduced by year ‘bubble’ crossover and non-compliance due forgetting and wanting to be ‘normal’ or socialise. Barriers to compliance noted in previous and our own data may be addressed through clear, consistent information and reminders, making things as easy as possible for families and staff, and engendering a sense of collective responsibility. Our data support the calls for extra government funding to support implementation of school mitigation measures.

The importance of clear and detailed guidance and school messaging which emphasises the collective good conflicts with a perceived lack of detail and clarity in the guidance issued by the UK government, and use of police and fines to enforce measures in wider society (51). }. Behavioural Scientists have expressed concern about this ‘blame’ approach and recommend instead an encouraging and compassionate approach which seeks to promoting collective identity and supportive social norms (52, 53). Schools will need to consider the benefits and risks of supportive rule enforcement vs a punitive approach, as well as potential stigma of Covid-19. Families and staff are generally accepting of school testing and contact tracing, but clear information about reporting symptoms (what, how, and why) and the use of personal data is needed.

Staff are concerned about the negative unintended consequences of school risk reduction measures, on behaviour, mental/emotional health, learning and pastoral care, particularly for SEN students and those disengaged from learning. Such concerns highlight the need for further research to identify the ongoing disproportionate impact of the Covid-19 pandemic on vulnerable and deprived groups, and the need for funding to provide additional learning and emotional support to students in need, There is also a need for research to understand how to address BAME families’ particular concerns relating to Covid-19 transmission within schools. Teachers’ wellbeing may also be impacted by feeling of guilt at possibility of infecting others, like other frontline workers.

### Limitations of this study

The main study limitation is limited inclusion of groups possibly most adversely affected, including: BAME/English as an additional language groups (although recruitment materials were translated, interviews were in English); those disengaged from school (as recruitment was mainly via schools); and those without internet access/computers as, due to Covid-19 restrictions we mainly used remote online recruitment (although we also used school newsletters and community groups). Also 19/20 parents were mothers.

## Conclusion

Families and staff supported Covid-19 mitigation measures in schools as a means of young people returning to face-to-face education. Clear messaging and engendering collective responsibility are important for measures’ compliance and success. However, schools and policy makers need to consider potential unintended consequences of measures, ways to support vulnerable individuals and those with additional needs, and how to avoid widening inequalities.

## Data Availability

The datasets used during the current study are available from the corresponding author on reasonable request.

## Acknowledgements

We would like to thank the valuable contribution of our participants and the community groups who helped with recruitment: Baggator; Barton Hill Activity Club; Black South West Network; Bristol Somali Voice; Bristol Somali Forum; BS5 Secondary Education Forum; The Care Forum; Felix Road Adventure Playground; Off The Record; St George Community Centre; Ujima Radio; Up Our Street; Young Bristol. We are also grateful to the ARC West YPAG and teachers who were involved in designing the study.

## Authorship

All authors have been involved in drafting the paper, read and approved the final version. AL recruited participants, collected and analysed data and led the writing of this manuscript. JK and JMK assisted with data collection. All authors collaborated on analysis.

